# Utilizing AI-Generated Plain Language Summaries to Enhance Interdisciplinary Understanding of Ophthalmology Notes: A Randomized Trial

**DOI:** 10.1101/2024.09.12.24313551

**Authors:** Prashant D. Tailor, Haley S. D’Souza, Clara M. Castillejo Becerra, Heidi M. Dahl, Neil R. Patel, Tyler M. Kaplan, Darrell Kohli, Erick D. Bothun, Brian G. Mohney, Andrea A. Tooley, Keith H. Baratz, Raymond Iezzi, Andrew J. Barkmeier, Sophie J. Bakri, Gavin W. Roddy, David Hodge, Arthur J. Sit, Matthew R. Starr, John J. Chen

## Abstract

**Background:** Specialized terminology employed by ophthalmologists creates a comprehension barrier for non-ophthalmology providers, compromising interdisciplinary communication and patient care. Current solutions such as manual note simplification are impractical or inadequate. Large language models (LLMs) present a potential low-burden approach to translating ophthalmology documentation into accessible language.

**Methods:** This prospective, randomized trial evaluated the addition of LLM-generated plain language summaries (PLSs) to standard ophthalmology notes (SONs). Participants included non-ophthalmology providers and ophthalmologists. The study assessed: (1) non-ophthalmology providers’ comprehension and satisfaction with either the SON (control) or SON+PLS (intervention), (2) ophthalmologists’ evaluation of PLS accuracy, safety, and time burden, and (3) objective semantic and linguistic quality of PLSs.

**Results:** 85% of non-ophthalmology providers (n=362, 33% response rate) preferred the PLS to SON. Non-ophthalmology providers reported enhanced diagnostic understanding (p=0.012), increased note detail satisfaction (p<0.001), and improved explanation clarity (p<0.001) for notes containing a PLS. The addition of a PLS narrowed comprehension gaps between providers who were comfortable and uncomfortable with ophthalmology terminology at baseline (intergroup difference p<0.001 to p>0.05). PLS semantic analysis demonstrated high meaning preservation (BERTScore mean F1 score: 0.85) with greater readability (Flesch Reading Ease: 51.8 vs. 43.6, Flesch-Kincaid Grade Level: 10.7 vs. 11.9). Ophthalmologists (n=489, 84% response rate) reported high PLS accuracy (90% “a great deal”) with minimal review time burden (94.9% ≤ 1 minute). PLS error rate on initial ophthalmologist review and editing was 26%, and 15% on independent ophthalmologist over-read of edited PLSs. 84.9% of identified errors were deemed low risk for patient harm and 0% had a risk of severe harm/death.

**Conclusions:** LLM-generated plain language summaries enhance accessibility and utility of ophthalmology notes for non-ophthalmology providers while maintaining high semantic fidelity and improving readability. PLS error rates underscore the need for careful implementation and ongoing safety monitoring in clinical practice.

## Introduction

The specialized nature and unique terminology of ophthalmology, combined with its limited coverage in medical education, create a comprehension gap for non-ophthalmology medical providers. This gap can hinder effective interdisciplinary communication and impact patient care. Current solutions, such as manual note simplification or abbreviation tables, are either impractical or insufficient.

Large Language Models (LLMs) offer a promising solution to this challenge. LLMs have demonstrated potential in various medical applications, including summarizing clinical notes, generating empathetic patient responses, and improving medical document readability (1–5). Prior studies exploring the usage of LLMs in medicine have largely been retrospective (5, 6). The real-world implementation of LLMs into clinical practice, particularly for improving interdisciplinary understanding, has to date been relatively unexplored.

The primary objectives of this study were to assess the efficacy of Plain Language Summaries (PLSs) in enhancing ophthalmology note comprehension among non-ophthalmology providers, evaluate the PLSs’ impact on ophthalmologists’ clinical workflow, and examine their accuracy and safety in real-world use.

## Methods

### Trial Design

This study was approved by the Mayo Clinic Institutional Review Board and adheres to the tenets of the Declaration of Helsinki. This trial was conducted as a quality improvement (QI) initiative.

This was a prospective, randomized controlled trial with two arms: a control arm with the ophthalmologist’s standard patient note (Standard Ophthalmology Note (SON)) only, and an intervention arm in which patient notes included a PLS appended to the end of the SON (**Figure 1**). Patient encounters were randomized to either the control or intervention arm following note completion to reduce bias. Three randomization schedules were utilized for different encounter types based on their location (inpatient, established outpatient, new outpatient consult) each with a block size of six.

**Figure 1.**
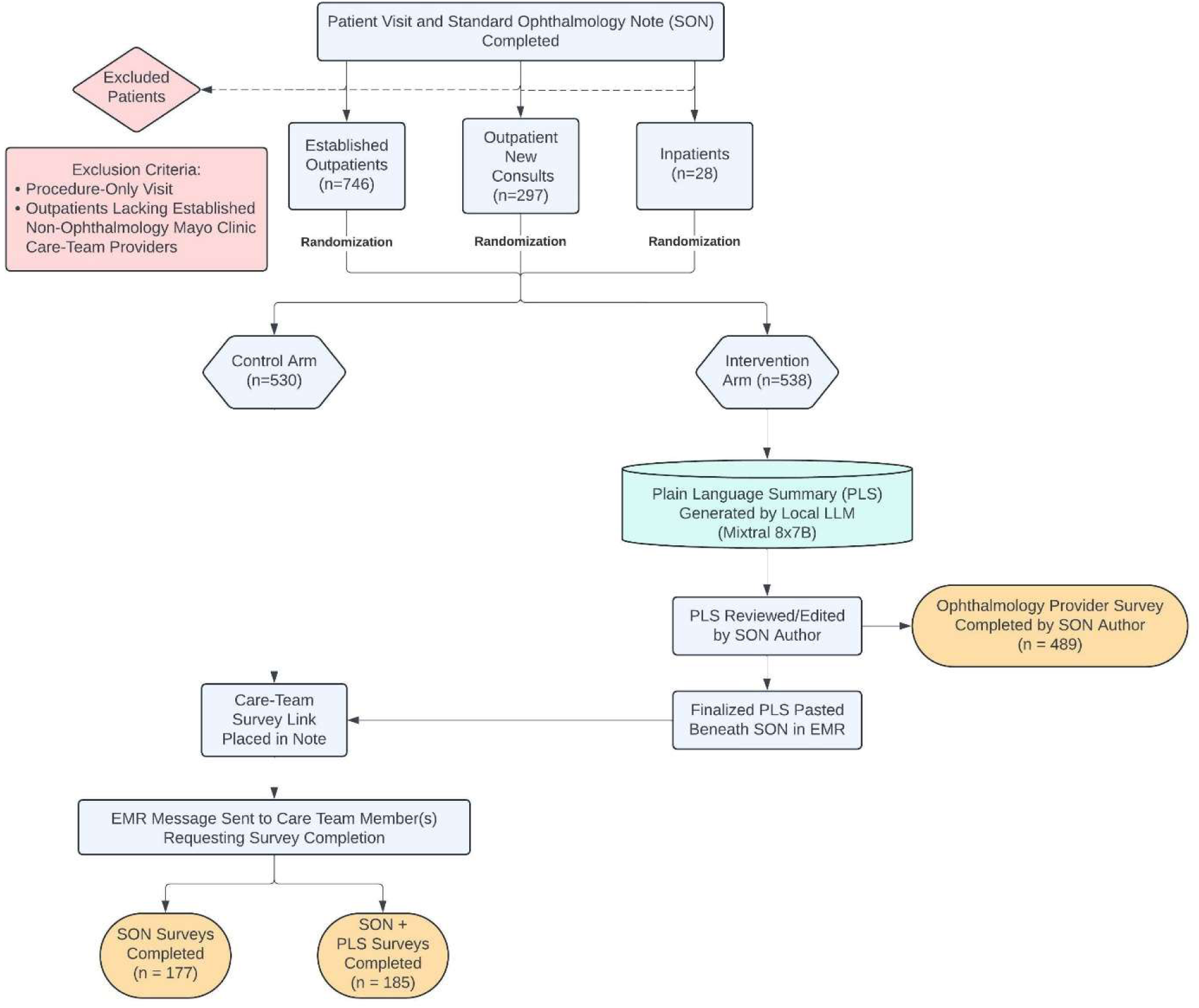
Study workflow for patient inclusion and exclusion, patient randomization, and control and intervention arm notes.

The primary endpoints were 1) Survey data from non-ophthalmology providers regarding clarity and readability of SON vs. SON with PLS, 2) Survey data from ophthalmologists regarding accuracy, time burden, and perception of PLSs, 3) Quantitative semantic and linguistic evaluation of PLSs, and 4) Safety analysis.

The single inclusion criterion was patient evaluation by an ophthalmologist in the Mayo Clinic Department of Ophthalmology in Rochester, Minnesota between February 1, 2024 and May 31, 2024. Exclusion criteria were: 1) Lack of established non-ophthalmology providers if the patient was seen in the outpatient setting, and 2) Procedure-only patient encounters (Supplementary Appendix: Exclusion Criteria).

### LLM Setup and Prompt Development

To ensure that protected health information was secure and to mitigate costs, Mixtral 8x7B (Mistral AI; Paris, France) was run locally on an encrypted 2021 Macbook Pro (M1 Max; 64GB). This model was selected as it was the top performing open-source model at the initiation of the study based on Elo rankings on the LMSYS.org Chatbot Arena Leaderboard. Prompt development was conducted prior to initiation of the study and the same prompt was used for the entirety of the study (Details in Supplementary Appendix; **Supplementary** Figure 1**, 2**).

### Study Work Flow

Following completion of a patient encounter, the encounter was randomized to either the control or intervention arm (**Figure 1**). For encounters randomized to the control arm, a survey link was placed at the end of the SON (**Supplementary** Figure 3) and was sent directly to non-ophthalmology care team provider(s) via standardized Electronic Medical Record (EMR) message (**Supplementary** Figure 4). If there were multiple members of the care team listed in the EMR, the survey was sent to each member (**Supplementary Table 1**).

For encounters randomized to the intervention arm, once the encounter was completed, a PLS was generated from the encounter’s SON using the LLM and prompt in a zero-shot fashion. The PLS was then reviewed (and edited if necessary) by the ophthalmologist who wrote the original SON. The ophthalmologist was instructed to edit the PLS solely for incorrect or missing information only. No stylistic changes (e.g. clarity, brevity, format etc.) were permitted. The ophthalmologist then completed a survey regarding the accuracy and burden of reading and, if necessary, editing the PLS (**Supplementary Table 2**). The finalized PLS was then inserted at the end of the SON along with the same survey link as the control arm (**Supplementary** Figure 5). An EMR message was sent to the non-ophthalmology provider(s) requesting their participation in completing the survey. If there were multiple members of the care team listed in the EMR, the survey was sent to each member. Neither the EMR message nor PLS disclosed that the PLS was generated by artificial intelligence to minimize respondent bias.

Surveys were designed with assistance from the Mayo Clinic Survey Research Center using Likert scales (Supplementary Appendix: Survey Design). Questions and scales were selected based on previously established frameworks for survey-based LLM evaluation.(6)

### Semantic and Lexical Evaluation

Standardized language metrics were used to assess the quality of PLSs versus SONs. This included readability (Flesch Reading Ease (FRE), Flesch-Kincaid Grade Level (FKGL), Simple Measure of Gobbledygook (SMOG) Index), semantic similarity (Bidirectional Encoder Representations from Transformers (BERT) Score, Sentence Transformers (SBERT) cosine similarity), and lexical overlap (BLEU-4, ROUGE-1) (Supplementary Appendix: Semantic Analysis).

### Patient Safety

To ensure patient safety, all PLSs were reviewed and edited to identify missing/inappropriate information by the ophthalmologist who wrote the original note prior to insertion of the PLS into the medical record. The finalized PLS was added to the end of the ophthalmology note following a clearly marked header (**Supplementary** Figure 5) to ensure it was easily delineated from the original note and to signify that the PLS was part of a QI initiative.

A random sample of PLSs and associated SONs were over-read by an independent ophthalmologist to evaluate for solely for inappropriate or missing information in the PLS not noted by the original author/reviewer. No stylistic issues were considered an error (e.g. clarity, brevity, format etc.). To reduce bias, this ophthalmologist did not contribute any SONs to the study. If any incorrect information in the PLS was identified following ophthalmologist overread, the inaccurate information was corrected in the EMR.

### Sample Size/Statistical Analyses

Using Cochran’s formula and based on a confidence level of 95%, a margin of error of 5% and an estimated population size of 5,000 non-ophthalmology providers at the Mayo Clinic in Rochester, Minnesota, the ideal sample size was determined to be 357 participants.

Data manipulation, numerical computations, visualizations and statistical analyses were performed using Python (version 3.9) and SAS (version 9.4) (Supplemental Appendix). Comparisons between groups for continuous variables were completed using Kruskal-Wallis tests. Categorical variables were compared using Chi-square tests. Potential relationships between continuous variables were assessed using Spearman’s correlation coefficients.

## Results

### Non-Ophthalmology Provider Survey

A total of 362 responses were collected (**Supplementary Table 3**). The intervention group (PLS + SON) consisted of 177 responses (48.9%), while the control group (SON only) had 185 responses (51.1%). The survey response rate was 32.8% and 34.7% for the intervention and control groups respectively. There were no significant differences between the control and intervention groups in specialty participation, role, and how often providers reviewed ophthalmology notes (**Supplementary Table 4**). Significant differences between the control and intervention groups were noted in baseline comfort with understanding ophthalmology notes and the impact of ophthalmology terminology on understanding of ophthalmology notes (**Figure 2**; p=0.04, p=0.03 respectively).

**Figure 2.**
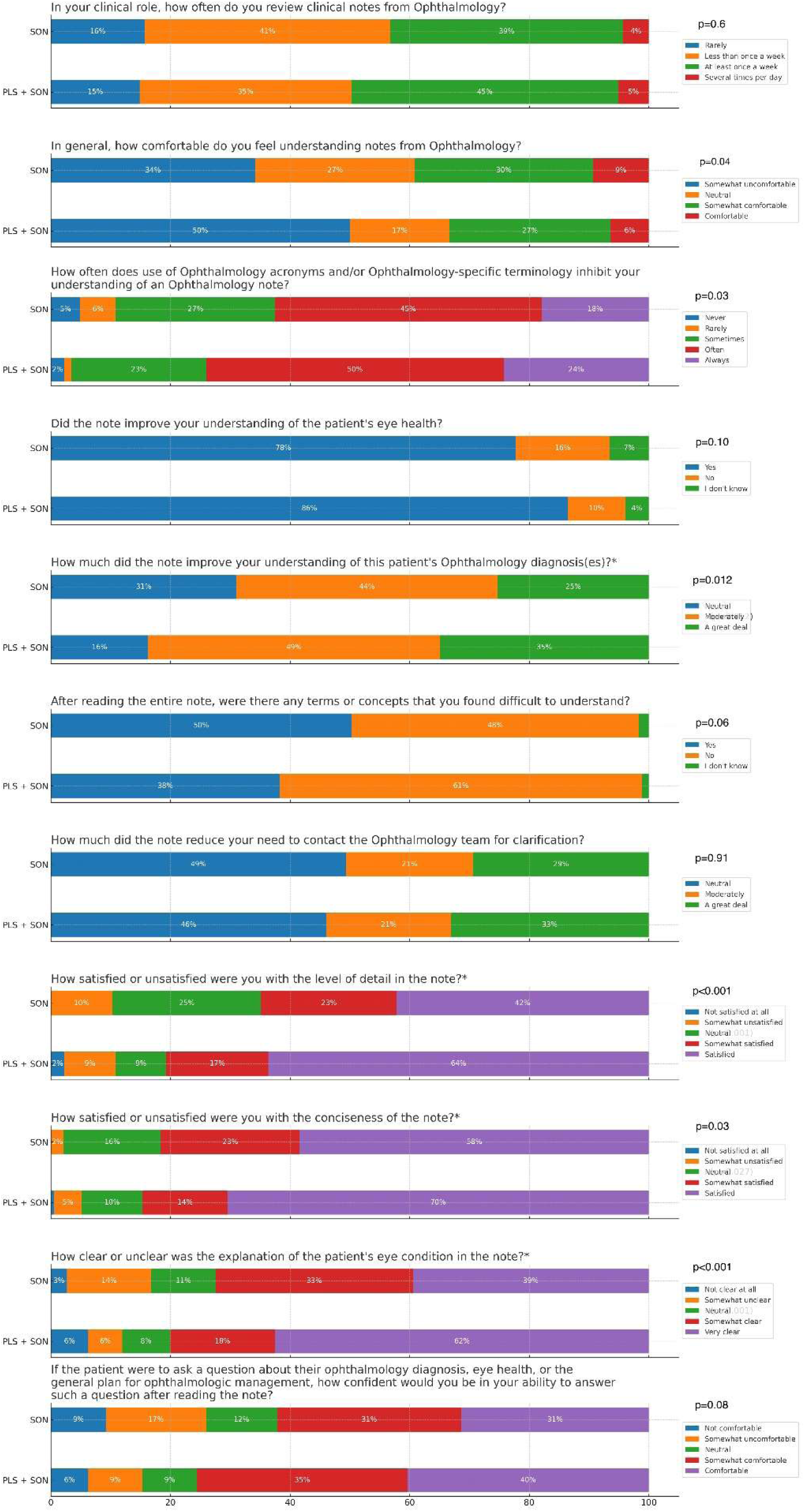
Non-ophthalmologist survey responses stratified by control and intervention.

The PLS+SON group was superior to the SON group for the following questions: 1. “How much did the note improve your understanding of this patient’s Ophthalmology diagnosis(es)?” (33% vs. 24% “a great deal”; p=0.012), 2. “How satisfied or unsatisfied were you with the level of detail in the note?” (64% vs. 42% “satisfied”; p<0.001), 3. “How satisfied or unsatisfied were you with the conciseness of the note?” (71% vs. 58% “satisfied”; p=0.027), and 4. “How clear or unclear was the explanation of the patient’s eye condition in the note?” (63% vs. 40%; “very clear”; p<0.001) (**Figure 2**).

When stratified by role, residents/fellows reported increased comfort in understanding and improved diagnosis comprehension (p=0.007 and p=0.047 respectively), while attending physicians reported higher satisfaction with note detail and clarity of explanation (**Supplementary Table 4**; p=0.025 and p=0.022 respectively).

The intervention group was asked three additional questions directly comparing the PLS to the SON (**Figure 3**). For the question “Which version provided clearer guidance regarding the patient’s treatment plan?” 85% preferred the PLS. For “Which version did you find easier to understand?” 88% preferred the PLS. Finally, when asked “Considering all aspects of your clinical practice, which type of note do you prefer?” 85% preferred the PLS over the SON. This preference was consistent across all non-ophthalmologist provider roles (**Supplementary Table 4**).

**Figure 3.**
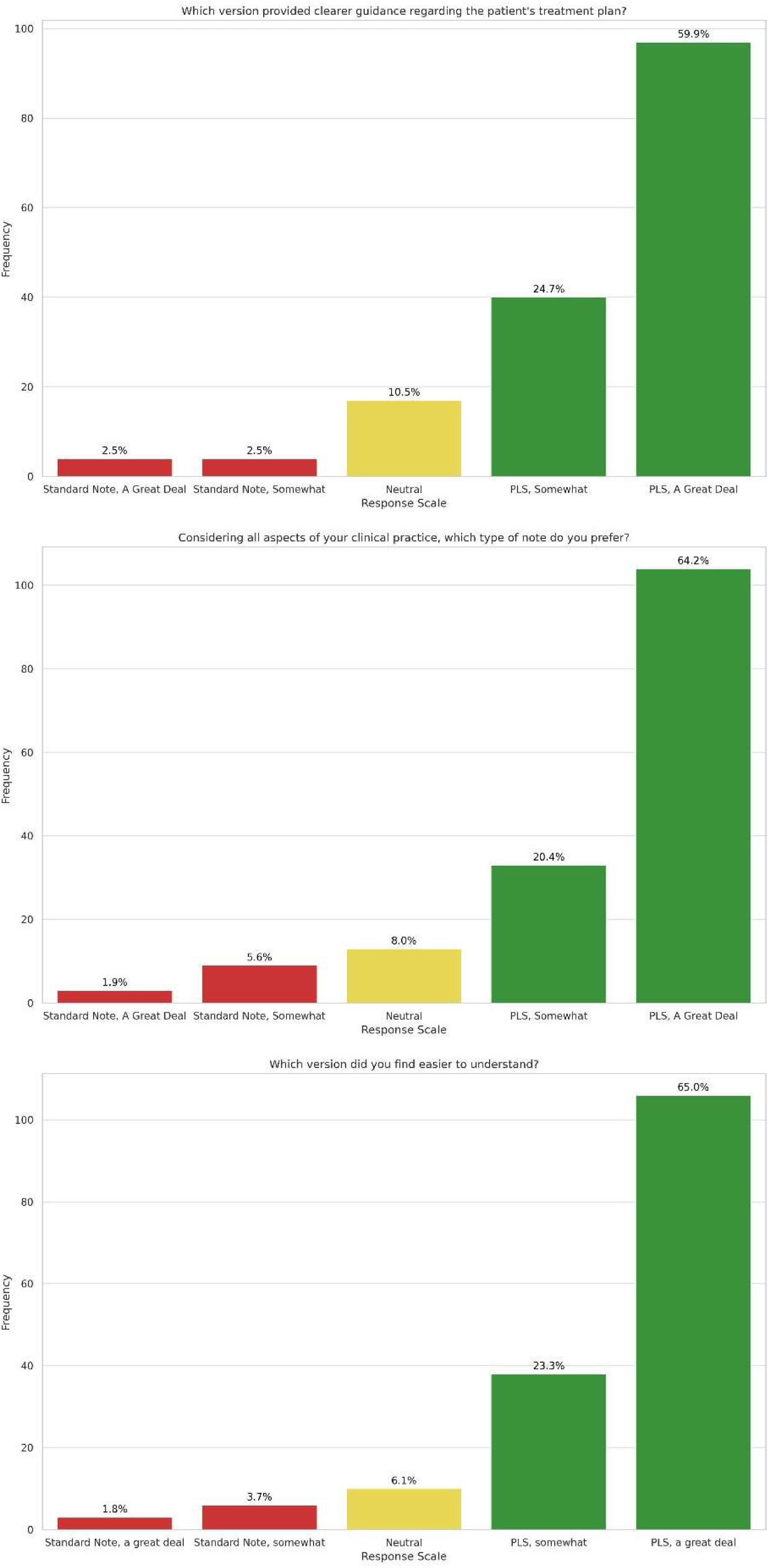
Intervention arm non-ophthalmologist survey responses comparing reader opinion of Standard Ophthalmology Note (SON) to Plain Language Summary (PLS).

Responses were stratified into Comfortable (Comfortable and Somewhat Comfortable) and Uncomfortable (Uncomfortable and Somewhat Uncomfortable) groups based on baseline comfort with reading ophthalmology notes **(Table 1)**. In the control group, 91.9% of Comfortable and 65.8% of Uncomfortable respondents reported improved understanding of the patient’s eye health (p=0.001). In the SON+PLS group, understanding was 94.2% and 79.8% respectively, with the difference being statistically insignificant (p=0.062). Significant differences in satisfaction with detail (61.9% Comfortable vs. 27.8% Uncomfortable; p<0.001) and clarity (55.6% Comfortable vs. 27.8% Uncomfortable; p<0.001) were observed in the control group, but the gap decreased with PLS addition, rendering the differences non-significant (Detail: 73.1% Comfortable vs. 60.6% Uncomfortable; p=0.070, Clarity: 67.3% Comfortable vs. 62.6% Uncomfortable; p=0.101). The gap in understanding due to unfamiliar terms also decreased in the intervention group but remained significant (control: 22.2% vs. 71.8%, p<0.001; intervention: 19.2% vs. 47.4%, p=0.002). Uncomfortable respondents showed a stronger preference for PLS over SON than Comfortable respondents (PLS a great deal: 75.6% Uncomfortable vs. 50.0% Comfortable; p=0.019).

**Table 1.**
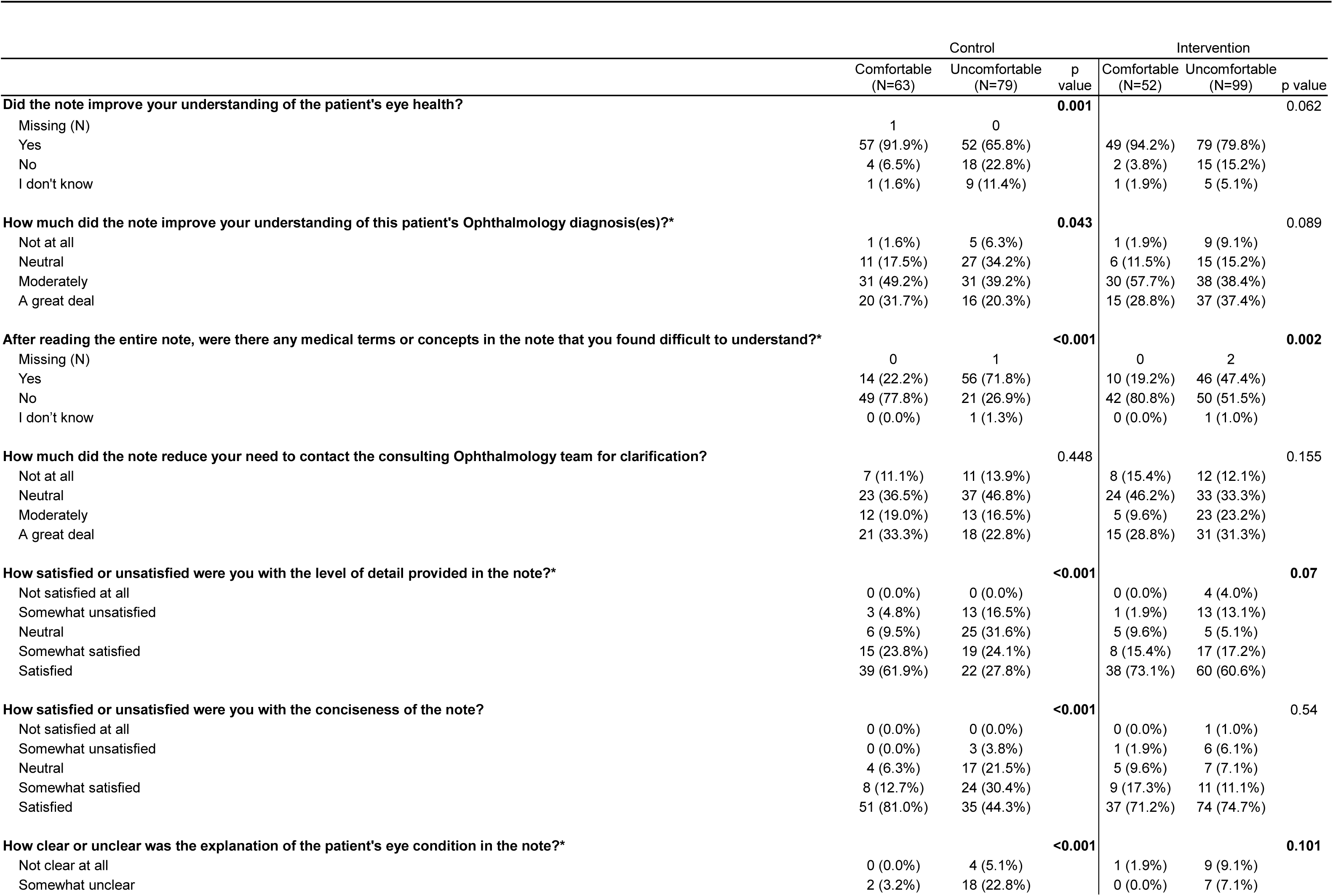
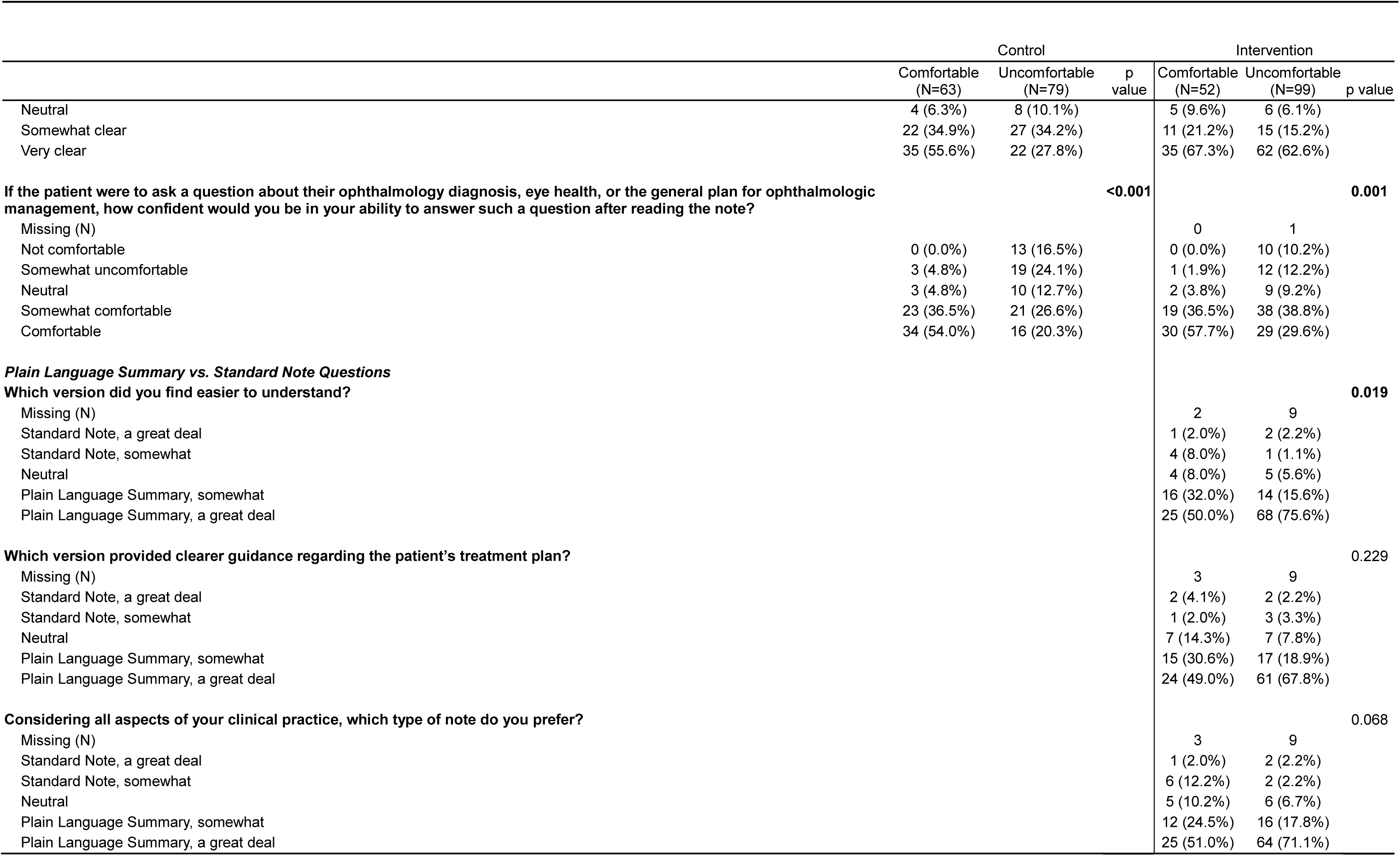
Comparison Between Intervention and Control Groups Stratified by Comfort with Ophthalmology Notes.

### Semantic and Lexical Similarity Evaluation

BERTScore demonstrated strong semantic correspondence between SONs and their PLSs, with mean scores of 0.85 (SD=0.04) for precision, 0.86 (SD=0.02) for recall, and 0.85 (SD=0.03) for F1. SBERT yielded a mean score of 0.84 (SD=0.06), indicating that PLS effectively preserved the meaning of the SON. Conversely, lexical overlap measures showed greater divergence between PLSs and their corresponding SONs, with a mean BLEU-4 score of 0.08 (SD=0.07) and a mean ROUGE-1 F-score of 0.47 (SD=0.11) (**Supplementary** Figure 6**, Supplementary Table 5**).

The mean FRE score significantly increased from a median of 43.6 (Interquartile Range (IQR) 19.8, range: -114.9 to 84.9) for SONs to a median of 51.8 (IQR=11.2, range: 17.3 to 70.4) for PLSs, indicating enhanced readability (p<0.001) (**Supplementary Table 6**). The FKGL decreased from a median of 11.9 (IQR=5.1, range: 4.4 to 77.0) for SONs to 10.7 (IQR=2.3, range: 6.5 to 22.0) for PLSs (p<0.001). The SMOG Index was unchanged in PLSs (median=13.0, IQR=1.9, range: 9.3 to 20.1) compared to SONs (median=13.0, IQR=3.6, range: 0.0 to 22.5), but with reduced variability (p=0.57).

### Ophthalmologist Survey

489 surveys (n=583; 84% response rate) were completed by ophthalmologists (**Table 2**). Respondents indicated that the PLS reflected the SON findings regarding diagnosis (90% ‘a great deal’) and plan (90.6% ‘a great deal’). 75.5% of respondents were ‘very satisfied’ with the PLS. Most respondents were ‘very satisfied’ with the PLS (attending physicians: 82.8%, trainees: 72.7%; p=0.001), but a larger proportion of trainees (20%) reported being ‘a little satisfied’ compared to 9.7% of attending physicians.

**Table 2.**
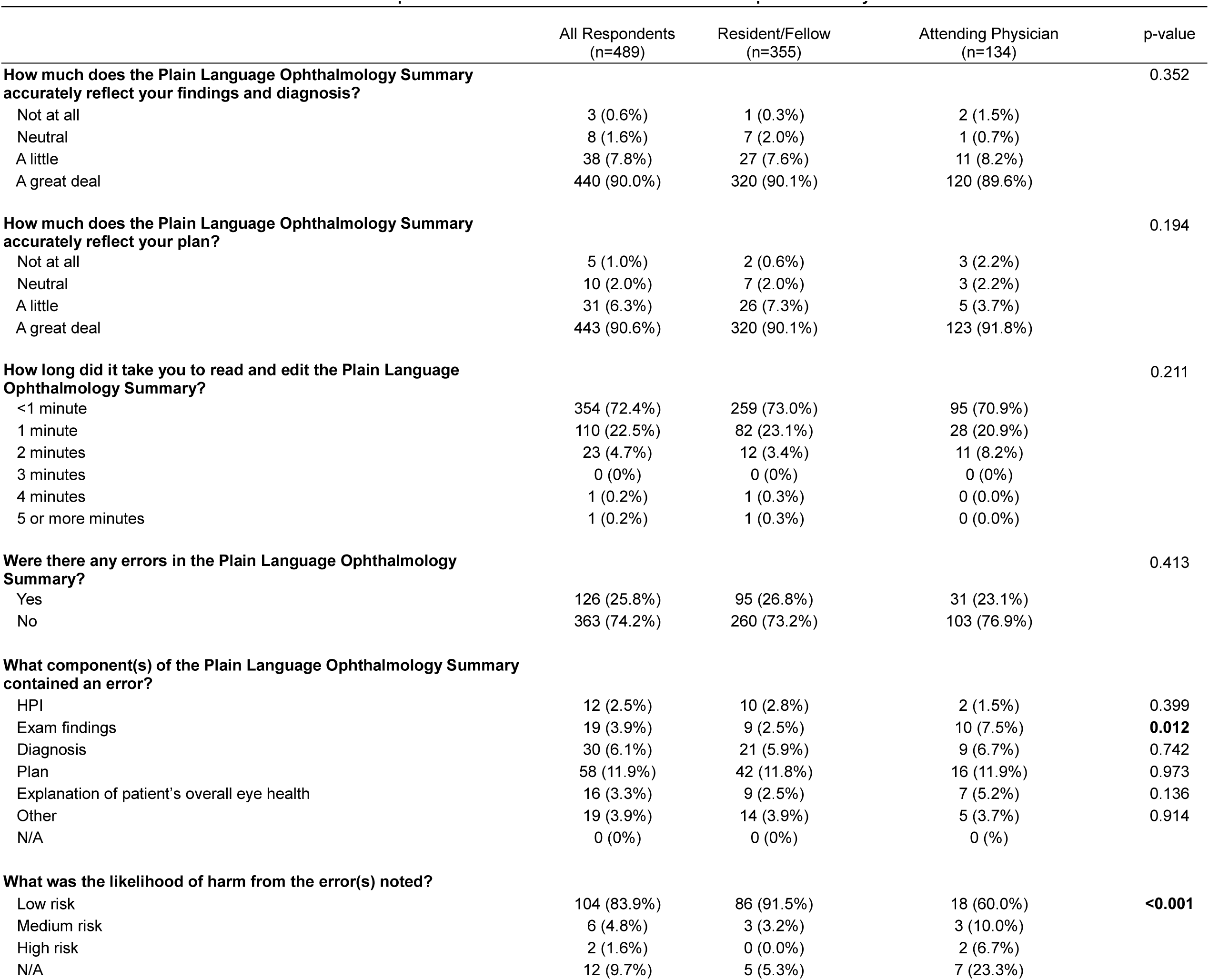
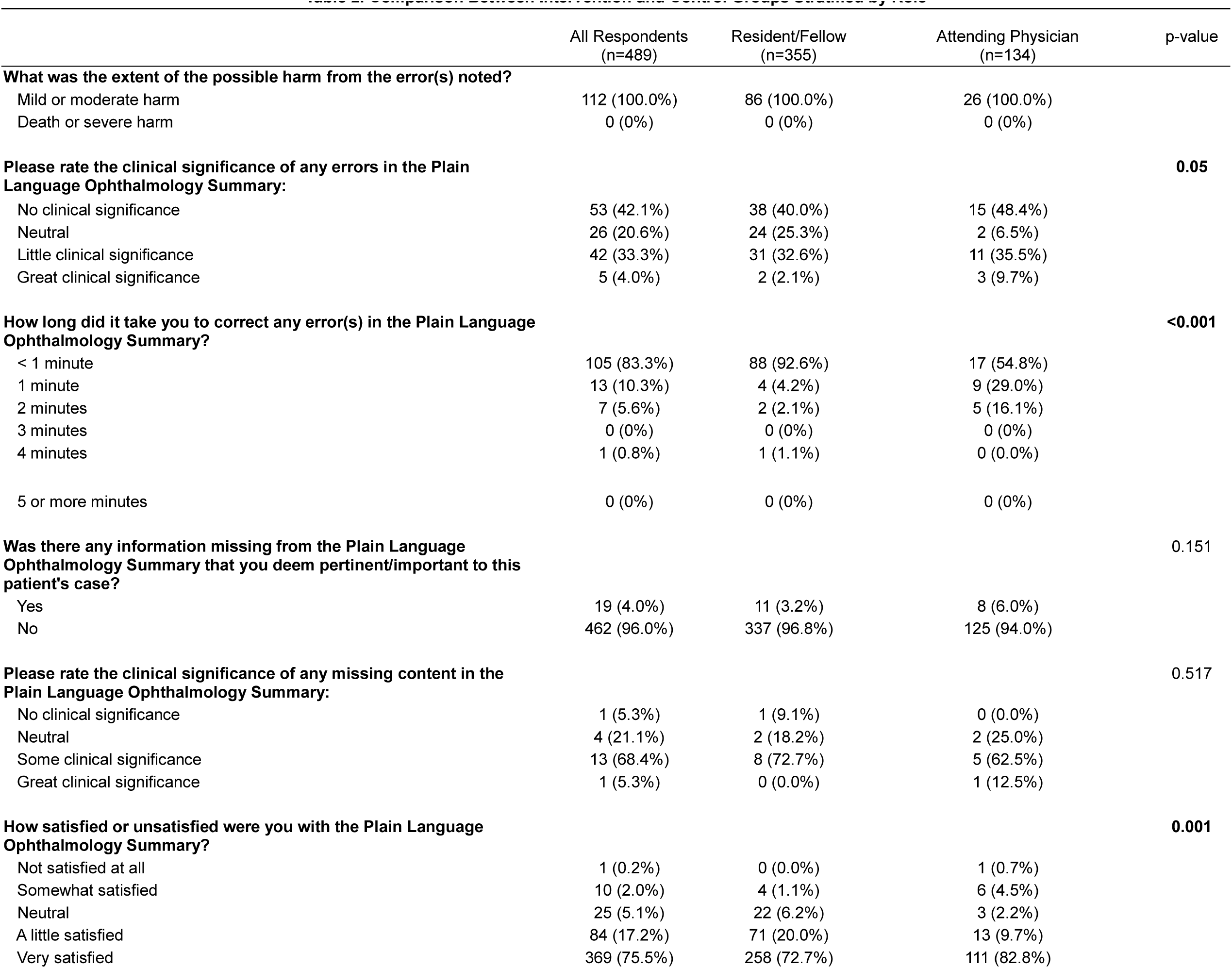
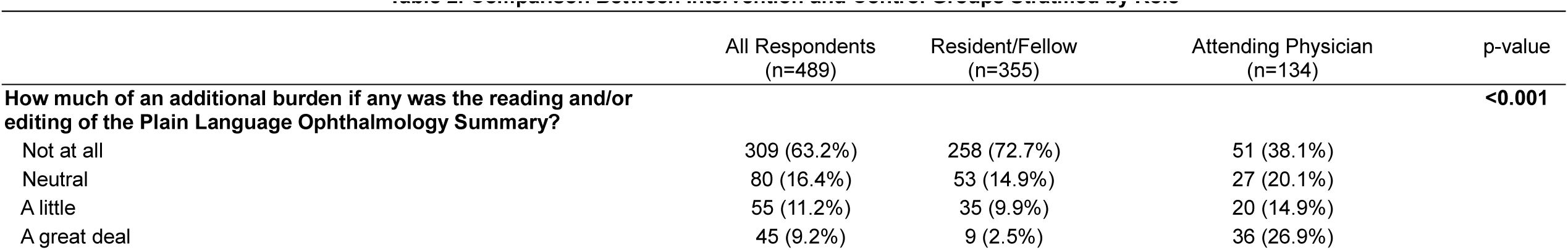
Comparison Between Intervention and Control Groups Stratified by Role.

72.4% of respondents spent <1 minute reading and editing the PLS. 63.2% of respondents reported that reading and/or editing the PLS was ‘not at all’ a burden, 16.4% ‘neutral,’ 11.2% ‘a little,’ and 9.2% ‘a great deal.’ There was a significant difference in the burden rating by attending physicians compared to trainees, with 26.9% (n=36) of attending physicians reporting ‘a great deal’ of burden compared to 2.5% (n=9) of trainees (p<0.001).

4.0% of PLSs were missing information (n=19) and 26% of PLSs contained inappropriate or incorrect information prior to editing. There was a significant difference in the proportion of attending physicians who reported an error in the examination findings (7.5%; n=10) compared to the proportion of trainees who reported an examination finding error (2.5%; n=9) (p=0.012).

83.9% of respondents indicated that the noted error(s) had low risk of harm, 4.8% medium risk, 1.6% high risk, and 9.7% were not applicable (no perceived risk of harm). There was a significant difference in the distribution of harm ratings by attending physicians vs. trainees, with 6.7% (n=2) of attending physicians reporting likelihood of ‘high risk’ and 0% of trainees reporting likelihood of ‘high risk’ of harm with the error(s) noted (p<0.001). All respondents indicated that the possibility of harm from any error(s) was ‘mild to moderate,’ and no respondents indicated that the possibility of harm was ‘death or severe harm.’

42.1% reported that the error(s) were of ‘no clinical significance,’ 20.6% ‘neutral,’ 33.3% ‘little clinical significance,’ and 4% ‘great clinical significance.’ 5.3% reported that any missing information was of ‘no clinical significance,’ 21.1% ‘neutral,’ 68.4% ‘some clinical significance,’ and 5.3% ‘great clinical significance.’ 83.3% of errors took <1 minute to correct. There was a significant difference in the error correction time between attending physicians vs. trainees, with 92.6% (n=88) of trainees reporting errors taking <1 minute to correct compared to 54.8% (n=17) of attending physicians (p<0.001).

There was no significant difference in the median SON word count between the control and intervention groups (p=0.92; median 167 and 167 respectively) or the number of diagnoses listed in the SON (p=0.54; median 4.0 and 4.0 respectively). There was no significant correlation between PLS word count and ophthalmology provider burden rating (p=0.384). Word count was not significantly correlated with responses to any questions comparing PLS to SON (Understanding p=0.69; Decision Making p=0.53; Overall p=0.55).

Overreads were completed for 235 intervention notes. There was a significant difference between SON word count (median 167.0, range: 25-629) and PLS word count (median 246, range: 61-600; p<0.001). PLS overread error rate was 15% (n=34) with no errors deemed significant enough for severe harm/death. PLSs with overread errors had significantly higher word counts (p=0.03; median 276.0 with errors vs. 241.0 without errors). No significant correlation was found between the time point in the study at which the PLS was generated and overread error rate (p=0.12).

## Discussion

The addition of LLM-generated PLSs to ophthalmology notes enhanced satisfaction and comprehension among non-ophthalmology providers with 85% overall preference for the PLS over SON. The use of PLS improved understanding of ophthalmology diagnoses, satisfaction with note detail, and clarity of explanations while accurately representing the SON with low review burden on ophthalmologists.

### Impact on Non-Ophthalmology Providers

The impact of the PLS varied across provider roles, with residents and fellows reporting increased comfort in understanding and improved diagnosis comprehension, while attending physicians noted higher satisfaction with note detail and clarity of explanations. This highlights the versatility of PLS in addressing diverse needs across experience levels in non-ophthalmology providers. The addition of a PLS also minimized the comprehension gap between providers who were comfortable and providers who were uncomfortable with ophthalmology terminology reflecting similar findings in consulting and in call centers with “low performers” (7, 8). Such equalization could be particularly beneficial in interdisciplinary settings, enhancing team-based care and reducing the impact of specialized knowledge on communication and care delivery.

### Semantic and Lexical Similarity Evaluation

Semantic and lexical similarity evaluations revealed that PLS maintained high semantic fidelity with the original notes while achieving effective linguistic simplification. High BERTScore metrics and SBERT scores indicated strong semantic correspondence and retained meaning, while lower BLEU-4 and ROUGE-1 scores suggested successful paraphrasing and simplification. Readability analyses supported the effectiveness of PLS, with improved FRE scores and reduced FKGLs, indicating that PLS required less advanced reading skills.

### Ophthalmologist Survey and Implications

Most ophthalmologists reported that the PLS accurately reflected their recorded findings, diagnoses, and plans. Overall satisfaction was high, but there was a significant difference in satisfaction levels between attending physicians and trainees. Attending physicians reported higher satisfaction but greater perceived burden than trainees (p=0.001 and p<0.001 respectively). Most attending and trainee respondents (94.9%) spent ≤1 minute reviewing and editing the PLS, suggesting efficient integration into workflow. These findings indicate that while AI-generated summaries are generally well-received, clinical implementation may need to be tailored to different experience levels and practice settings to minimize burden. The perceived burden of AI integration, particularly among attending physicians, indicates a need for thoughtful integration into current workflows.

Errors were noted in 26% of PLSs, with the plan section being the most error-prone. Attending physicians detected more errors in examination findings compared to trainees. While most errors (83.3%) took less than one minute to correct, attending physicians reported longer correction times compared to trainees (p<0.001). 83.9% of these errors were considered to have a low risk of harm. Although no errors were deemed to have death or severe harm’ harm risk, the 15% overread error rate raises concerns about the need for oversight of AI tools to ensure patient safety, especially given increasing clinical volumes.

### Limitations

This study has several limitations. The use of survey data introduces subjectivity and potential response bias. Clear PLS labeling as QI may have biased non-ophthalmology providers to review the PLS favorably in comparison with the SON. Ideally, providers would receive either SON or PLS randomly, but this requires further rigorous safety evaluation and testing due to placing AI generated information in the medical record without clear labelling. The use of Mixtral 7x8B, no longer the top-performing model, indicates that results may not reflect the capabilities current state-of-the-art models. Additionally, this study does not directly measure impact on patient outcomes or long-term effects on interdisciplinary collaboration. Finally, this was a single-site study limited to a single tertiary academic institution and may not generalize to other settings. Despite these limitations, the statistically significant findings and demonstrated improvements in non-ophthalmologist comprehension and satisfaction provide a strong foundation for future research.

## Conclusion

The addition of LLM-generated plain language summaries to ophthalmology notes significantly improves comprehension and satisfaction among non-ophthalmology providers, with overall satisfaction and minimal time burden for ophthalmologists in addition to objective improvement in readability. Plain language summaries have the potential to enhance interdisciplinary communication, reduce cognitive load, and make medical information more accessible, improving patient care. Future research should test LLM-generated summaries in other specialties and settings to validate this approach and advance AI tools in healthcare. The presence of errors in AI-generated summaries despite human oversight highlights the need for careful implementation and ongoing safety monitoring to balance improved communication with patient safety.

## Data Availability

All data produced in the present work are contained in the manuscript

