## Supplementary Appendix for "Utilizing AI-Generated Plain Language Summaries to Enhance Interdisciplinary Understanding of Ophthalmology Notes: A Randomized Trial"

**Table of Contents**

1. Supplementary Methods…………………………………………….. Page 2
2. Supplementary Tables……………………………………………….. Page 4
3. Supplementary Figures………………………………………………. Page 18

This appendix has been provided by the authors to give readers additional information about their work.

Supplement to: Tailor PD, D'Souza HS, Castillejo Becerra C, Dahl HM, Patel NR, Kaplan TM, et al. Utilizing AI-Generated Plain Language Summaries to Enhance Interdisciplinary Understanding of Ophthalmology Notes: A Randomized Trial.

### Supplementary Methods

*Exclusion Criteria*

For the purposes of this study, an established outpatient non-ophthalmology provider was defined as any provider within the Mayo Clinic network in a non-ophthalmology specialty who had seen the patient over two or more patient encounters within the last two years, a referring provider, or someone designated in the electronic medical record (EMR) as either the patient's primary care provider or member of the patient's care team. Inpatient non-ophthalmology providers were defined as any members of the healthcare team listed in the patient's care team within the EMR. Patients without established non-ophthalmology providers within the Mayo Clinic network were excluded as there was no reliable mechanism to obtain non-Ophthalmologist survey feedback.

*LLM Prompt Development*

Prompt development was conducted prior to study initiation in an iterative fashion utilizing real patient encounters at all encounter types (inpatient, established outpatient, and new outpatient consult) in all ophthalmology subspecialties: cornea, glaucoma, uveitis, retina, oculoplastics, neuro-ophthalmology, cataract/refractive, and pediatric ophthalmology. Modifications to the prompt during development were based on expert reviews by two ophthalmologists of LLM outputs. The aim was to ensure that the output was free of jargon, provided explanations of subspecialty testing and diagnoses, and clearly stated a plan and follow-up. We chose a prompt-based approach over a fine-tuned LLM for several reasons: 1) prompt modification is easy to implement, deploy, and refine, allowing quick adjustments without extensive retraining; 2) it is more cost-effective, reducing the need for significant computational resources; and 3) it enhances the model's generalizability, making it easier for other researchers to replicate and apply, fostering broader adoption. Once finalized, the prompt (**Supplementary Figure 1**) was not modified during the entire study period. The model parameter settings were fixed to the default for the entire study period (**Supplementary Figure 2**).

*Survey Design*

Surveys were designed using Likert scales for reliability and validity with the Mayo Clinic Survey Research Center. Questions and scales were selected based on previously established frameworks for survey-based LLM evaluation.(1)

The non-ophthalmology provider survey (**Supplementary Table 2**) assessed three primary areas of interest: 1) The responder's role and subspecialty, 2) Prior experience/comfort with reading ophthalmology notes, and 3) Understanding of the note in terms of the diagnosis, plan, terminology, level of detail, and explanation of eye health. If the provider indicated that they had viewed a note containing a PLS, the survey also assessed for a direct comparison of the SON to the PLS with regards to clarity, guidance in the treatment plan, and overall preference.

The ophthalmologist survey evaluated ophthalmologists' experiences reading and editing the PLS (**Supplementary Table 1**). This survey appraised four primary areas of interest: 1) Time taken to read the PLS and edit if necessary, 2) PLS accuracy (in terms of diagnosis and plan) 3) Type of errors identified and their possible harm, and 4) Perceived additional burden from reading and editing. The questions and scales on inappropriate content, missing content, extent of possible harm, and likelihood of possible harm were adapted from Singhal et al.(1)

*Semantic Analysis*

To objectively evaluate the quality of PLSs in relation to their corresponding SON, we employed a multifaceted approach using standardized language evaluation metrics. Readability was assessed using three indices: Flesch Reading Ease, Flesch-Kincaid Grade Level (FKGL), and Simple Measure of Gobbledygook (SMOG) Index, providing a comprehensive measure of linguistic accessibility. Semantic similarity between each PLS and its corresponding SON was evaluated using two natural language processing techniques: Bidirectional Encoder Representations from Transformers (BERT) Score, which provides precision, recall, and F1 scores based on contextual embeddings, and Sentence Transformers (SBERT) with cosine similarity, which provides a holistic measure of overall semantic correspondence. Additionally, lexical overlap was quantified using Bilingual Evaluation Understudy (BLEU-4) and Recall-Oriented Understudy for Gisting Evaluation (ROUGE-1), assessing n-gram precision and unigram recall, respectively. The NTLK library was used for general text processing and BLEU score calculation, the rouge_score library was utilized to compute ROUGE-1 scores, and the bert_score package was used to calculate BERTScores. For sentence embeddings and cosine similarity calculations, the sentence-transformers library, specifically the "all-mpnet-base-v2" model for Sentence-BERT (SBERT) computations, was utilized. Readability metrics were calculated using the textstat library.

### Supplementary Tables

**Supplementary Table 1.** Non-ophthalmology provider survey questions.

**Questions answered by all respondents:**

1. **Patient MRN**
2. **Please select your specialty:**
    a. Internal Medicine / Pediatrics / Medicine Subspecialty
    b. Surgery / Surgical Subspecialty
    c. Emergency Medicine
    d. Other
    e. Family Medicine
3. **Your Role**
    a. Attending
    b. Fellow
    c. Resident
    d. Medical Student
    e. Pharmacist
    f. PA
    g. NP
    h. Nurse
    i. Other
4. **In your clinical role, how often do you review clinical notes from Ophthalmology?**
    a. Rarely
    b. Less than once a week
    c. At least once a week
    d. Several times per day
5. **In general, how comfortable do you feel understanding notes from Ophthalmology?**
    a. Not comfortable
    b. Somewhat uncomfortable
    c. Neutral
    d. Somewhat comfortable
    e. Comfortable
6. **How often does use of Ophthalmology acronyms and/or Ophthalmology-specific terminology inhibit your understanding of an Ophthalmology note (i.e. APD, LPI, SLT, YAG)?**
    a. Never
    b. Rarely
    c. Sometimes
    d. Often
    e. Always
7. **Did the note improve your understanding of the patient's eye health?**
    a. Yes
    b. No
    c. I don't know
8. **How much did the note improve your understanding of this patient's Ophthalmology diagnosis(es)?**
    a. Not at all
    b. Neutral
    c. Moderately
    d. A great deal
9. **After reading the entire note, were there any medical terms or concepts in the note that your found difficult to understand?**
    a. Yes
    b. No

c. I don’t know

10. **How much did the note reduce your need to contact the consulting Ophthalmology team for clarification?**
    a. Not at all
    b. Neutral
    c. Moderately
    d. A great deal
11. **How satisfied or unsatisfied were you with the level of detail provided in the note?**
    a. Not satisfied at all
    b. Somewhat unsatisfied
    c. Neutral
    d. Somewhat satisfied
    e. Satisfied
12. **How satisfied or unsatisfied were you with the conciseness of the note?**
    a. Not satisfied at all
    b. Somewhat unsatisfied
    c. Neutral
    d. Somewhat satisfied
    e. Satisfied
13. **How clear or unclear was the explanation of the patient's eye condition in the note?**
    a. Not clear at all
    b. Somewhat unclear
    c. Neutral
    d. Somewhat clear
    e. Very clear
14. **If the patient were to ask a question about their ophthalmology diagnosis, eye health, or the general plan for ophthalmologic management, how confident would you be in your ability to answer such a question after reading the note?**
    a. Not comfortable
    b. Somewhat uncomfortable
    c. Neutral
    d. Somewhat comfortable
    e. Comfortable
15. **Did you view a note containing a Plain Language Ophthalmology Summary? (see above photo for example)**
    a. Yes
    b. No
16. **Please add any additional comments or feedback for improvement of the Plain Language Ophthalmology Summary here:** [free text response]

---

**Questions answered by respondents in intervention arm:**

17. **Which version did you find easier to understand?**
    a. Standard Note, a great deal
    b. Standard Note, somewhat
    c. Neutral
    d. Plain Language Summary, somewhat
    e. Plain Language Summary, a great deal
18. **Which version provided clearer guidance regarding the patient's treatment plan?**
    a. Standard Note, a great deal
    b. Standard Note, somewhat
    c. Neutral
    d. Plain Language Summary, somewhat
    e. Plain Language Summary, a great deal
19. **Considering all aspects of your clinical practice, which type of note do you prefer?**
    a. Standard Note, a great deal
    b. Standard Note, somewhat

c. Neutral
d. Plain Language Summary, somewhat
e. Plain Language Summary, a great deal

**Supplementary Table 2.** Ophthalmologist survey questions.

1. Patient MRN
2. Your Name
3. Your Role
    a. Attending
    b. Fellow
    c. Resident
    d. Other
4. How much does the Plain Language Ophthalmology Summary accurately reflect your findings and diagnosis?
    a. Not at all
    b. Neutral
    c. A little
    d. A great deal
5. How much does the Plain Language Ophthalmology Summary accurately reflect your plan?
    a. Not at all
    b. Neutral
    c. A little
    d. A great deal
6. How long did it take you to read and edit the Plain Language Ophthalmology Summary?
    a. < 1 minute
    b. 1 minute
    c. 2 minutes
    d. 3 minutes
    e. 4 minutes
    f. 5 or more minutes
7. Were there any errors in the Plain Language Ophthalmology Summary?
    a. Yes
    b. No
8. Was there any information missing from the Plain Language Ophthalmology Summary that you deem pertinent/important to this patient's case?
    a. Yes
    b. No
9. How satisfied or unsatisfied were you with the Plain Language Ophthalmology Summary?
    a. Not satisfied at all
    b. Somewhat unsatisfied
    c. Neutral
    d. A little satisfied
    e. Very satisfied
10. How much of an additional burden if any was the reading and/or editing of the Plain Language Ophthalmology Summary?
    a. Not at all
    b. Neutral
    c. A little
    d. A great deal
11. Please add any additional comments or feedback for improvement of the Plain Language Ophthalmology Summary here: [free text response]

Questions answered by respondents if ‘Yes’ to Question 7:

12. What component(s) of the Plain Language Ophthalmology Summary contained an error?
    a. HPI
    b. Exam findings
    c. Diagnosis
    d. Plan

    e. Explanation of patient's overall eye health
    f. Other
    g. N/A
13. What was the likelihood of harm from the error(s) noted?
    a. Low risk
    b. Medium risk
    c. High risk
    d. N/A
14. What was the extent of the possible harm from the error(s) noted?
    a. Mild or moderate harm
    b. Death or severe harm
15. Please rate the clinical significance of any errors in the Plain Language Ophthalmology Summary:
    a. No clinical significance
    b. Neutral
    c. Little clinical significance
    d. Great clinical significance
16. How long did it take you to correct any error(s) in the Plain Language Ophthalmology Summary?
    a. < 1 minute
    b. 1 minute
    c. 2 minutes
    d. 3 minutes
    e. 4 minutes
    f. 5 or more minutes

Question answered by respondents if 'Yes' to Question 8:

17. Please rate the clinical significance of any missing content in the Plain Language Ophthalmology Summary:
    a. No clinical significance
    b. Neutral
    c. Some clinical significance
    d. Great clinical significance

**Supplementary Table 3.** Non-ophthalmologist survey responses stratified by control and treatment groups.

| | Intervention (N=177) | Control (N=185) | p value |
|---|---|---|---|
| **In your clinical role, how often do you review clinical notes from Ophthalmology?** | | | 0.658 |
| Missing | 2 | 0 | |
| Rarely | 26 (14.9%) | 29 (15.7%) | |
| Less than once a week | 62 (35.4%) | 76 (41.1%) | |
| At least once a week | 78 (44.6%) | 72 (38.9%) | |
| Several times per day | 9 (5.1%) | 8 (4.3%) | |
| **In general, how comfortable do you feel understanding notes from Ophthalmology?** | | | **0.048** |
| Not comfortable | 21 (11.9%) | 24 (13.0%) | |
| Somewhat uncomfortable | 78 (44.1%) | 55 (29.7%) | |
| Neutral | 26 (14.7%) | 43 (23.2%) | |
| Somewhat comfortable | 42 (23.7%) | 48 (25.9%) | |
| Comfortable | 10 (5.6%) | 15 (8.1%) | |
| **How often does use of Ophthalmology acronyms and/or Ophthalmology-specific terminology inhibit your understanding of an Ophthalmology note (i.e. APD, LPI, SLT, YAG)?** | | | **0.033** |
| Missing | 0 | 1 | |
| Never | 4 (2.3%) | 9 (4.9%) | |
| Rarely | 2 (1.1%) | 11 (6.0%) | |
| Sometimes | 40 (22.6%) | 49 (26.6%) | |
| Often | 88 (49.7%) | 82 (44.6%) | |
| Always | 43 (24.3%) | 33 (17.9%) | |
| **Did the note improve your understanding of the patient's eye health?** | | | 0.098 |
| Missing | 0 | 1 | |
| Yes | 153 (86.4%) | 143 (77.7%) | |
| No | 17 (9.6%) | 29 (15.8%) | |
| I don't know | 7 (4.0%) | 12 (6.5%) | |
| **How much did the note improve your understanding of this patient’s Ophthalmology diagnosis(es)?** | | | **0.012** |
| Not at all | 11 (6.2%) | 11 (5.9%) | |
| Neutral | 27 (15.3%) | 54 (29.2%) | |
| Moderately | 81 (45.8%) | 76 (41.1%) | |

| | Intervention (N=177) | Control (N=185) | p value |
|---|---|---|---|
| A great deal | 58 (32.8%) | 44 (23.8%) | |
| **After reading the entire note, were there any medical terms or concepts in the note that your found difficult to understand?** | | | 0.060 |
| Missing | 2 | 2 | |
| Yes | 67 (38.3%) | 92 (50.3%) | |
| No | 106 (60.6%) | 88 (48.1%) | |
| I don't know | 2 (1.1%) | 3 (1.6%) | |
| **How much did the note reduce your need to contact the consulting Ophthalmology team for clarification?** | | | 0.906 |
| Not at all | 23 (13.0%) | 25 (13.5%) | |
| Neutral | 71 (40.1%) | 79 (42.7%) | |
| Moderately | 32 (18.1%) | 34 (18.4%) | |
| A great deal | 51 (28.8%) | 47 (25.4%) | |
| **How satisfied or unsatisfied were you with the level of detail provided in the note?** | | | **<0.001** |
| Missing | 1 | 0 | |
| Not satisfied at all | 4 (2.3%) | 0 (0.0%) | |
| Somewhat unsatisfied | 15 (8.5%) | 19 (10.3%) | |
| Neutral | 15 (8.5%) | 46 (24.9%) | |
| Somewhat satisfied | 30 (17.0%) | 42 (22.7%) | |
| Satisfied | 112 (63.6%) | 78 (42.2%) | |
| **How satisfied or unsatisfied were you with the conciseness of the note?** | | | **0.027** |
| Missing | 1 | 0 | |
| Not satisfied at all | 1 (0.6%) | 0 (0.0%) | |
| Somewhat unsatisfied | 8 (4.5%) | 4 (2.2%) | |
| Neutral | 18 (10.2%) | 30 (16.2%) | |
| Somewhat satisfied | 25 (14.2%) | 43 (23.2%) | |
| Satisfied | 124 (70.5%) | 108 (58.4%) | |
| **How clear or unclear was the explanation of the patient's eye condition in the note?** | | | **<0.001** |
| Missing | 1 | 0 | |
| Not clear at all | 11 (6.3%) | 5 (2.7%) | |
| Somewhat unclear | 10 (5.7%) | 26 (14.1%) | |
| Neutral | 14 (8.0%) | 20 (10.8%) | |

| | Intervention (N=177) | Control (N=185) | p value |
|---|---|---|---|
| Somewhat clear | 31 (17.6%) | 61 (33.0%) | |
| Very clear | 110 (62.5%) | 73 (39.5%) | |
| **If the patient were to ask a question about their ophthalmology diagnosis, eye health, or the general plan for ophthalmologic management, how confident would you be in your ability to answer such a question after reading the note?** | | | 0.080 |
| Missing | 1 | 0 | |
| Not comfortable | 11 (6.3%) | 17 (9.2%) | |
| Somewhat uncomfortable | 16 (9.1%) | 31 (16.8%) | |
| Neutral | 16 (9.1%) | 22 (11.9%) | |
| Somewhat comfortable | 62 (35.2%) | 57 (30.8%) | |
| Comfortable | 71 (40.3%) | 58 (31.4%) | |
| **Did you view a note containing a Plain Language Ophthalmology Summary? (see above photo for example)** | | | |
| Yes | 165 (94.3%) | | |
| **Which version did you find easier to understand?** | | | |
| Missing | 14 | | |
| Standard Note, a great deal | 3 (1.8%) | | |
| Standard Note, somewhat | 6 (3.7%) | | |
| Neutral | 10 (6.1%) | | |
| Plain Language Summary, somewhat | 38 (23.3%) | | |
| Plain Language Summary, a great deal | 106 (65.0%) | | |
| **Which version provided clearer guidance regarding the patient's treatment plan?** | | | |
| Missing | 15 | | |
| Standard Note, a great deal | 4 (2.5%) | | |
| Standard Note, somewhat | 4 (2.5%) | | |
| Neutral | 17 (10.5%) | | |
| Plain Language Summary, somewhat | 40 (24.7%) | | |
| Plain Language Summary, a great deal | 97 (59.9%) | | |
| **Considering all aspects of your clinical practice, which type of note do you prefer?** | | | |
| Missing | 15 | | |
| Standard Note, a great deal | 3 (1.9%) | | |

| | Intervention (N=177) | Control (N=185) | p value |
|---|---|---|---|
| Standard Note, somewhat | 9 (5.6%) | | |
| Neutral | 13 (8.0%) | | |
| Plain Language Summary, somewhat | 33 (20.4%) | | |
| Plain Language Summary, a great deal | 104 (64.2%) | | |

**Supplementary Table 4.** Non-ophthalmologist survey responses stratified by role of the respondents.

| | Resident/Fellow | | | Staff Physician | | | NP/PA | | | Allied Health | | |
|---|---|---|---|---|---|---|---|---|---|---|---|---|
| | Intervention (N=22) | Control (N=37) | p value | Intervention (N=87) | Control (N=91) | p value | Intervention (N=47) | Control (N=38) | p value | Intervention (N=21) | Control (N=19) | p value |
| **In your clinical role, how often do you review clinical notes from Ophthalmology?** | | | 0.657 | | | 0.752 | | | 0.903 | | | 0.609 |
| Missing (N) | | | | 2 | 0 | | | | | | | |
| Rarely | 3 (13.6%) | 6 (16.2%) | | 7 (8.2%) | 8 (8.8%) | | 6 (12.8%) | 5 (13.2%) | | 10 (47.6%) | 10 (52.6%) | |
| Less than once a week | 14 (63.6%) | 26 (70.3%) | | 22 (25.9%) | 30 (33.0%) | | 21 (44.7%) | 14 (36.8%) | | 5 (23.8%) | 6 (31.6%) | |
| At least once a week | 5 (22.7%) | 5 (13.5%) | | 48 (56.5%) | 46 (50.5%) | | 19 (40.4%) | 18 (47.4%) | | 6 (28.6%) | 3 (15.8%) | |
| Several times per day | 0 (0%) | 0 (0%) | | 8 (9.4%) | 7 (7.7%) | | 1 (2.1%) | 1 (2.6%) | | 0 (0.0%) | 0 (0.0%) | |
| **In general, how comfortable do you feel understanding notes from Ophthalmology?*** | | | **0.007** | | | 0.233 | | | 0.107 | | | 0.207 |
| Not comfortable | 3 (13.6%) | 10 (27.0%) | | 5 (5.7%) | 8 (8.8%) | | 7 (14.9%) | 3 (7.9%) | | 6 (28.6%) | 3 (15.8%) | |
| Somewhat uncomfortable | 16 (72.7%) | 10 (27.0%) | | 42 (48.3%) | 31 (34.1%) | | 12 (25.5%) | 11 (28.9%) | | 8 (38.1%) | 3 (15.8%) | |
| Neutral | 2 (9.1%) | 7 (18.9%) | | 14 (16.1%) | 24 (26.4%) | | 7 (14.9%) | 5 (13.2%) | | 3 (14.3%) | 7 (36.8%) | |
| Somewhat comfortable | 0 (0.0%) | 9 (24.3%) | | 18 (20.7%) | 22 (24.2%) | | 20 (42.6%) | 12 (31.6%) | | 4 (19.0%) | 5 (26.3%) | |
| Comfortable | 1 (4.5%) | 1 (2.7%) | | 8 (9.2%) | 6 (6.6%) | | 1 (2.1%) | 7 (18.4%) | | 0 (0.0%) | 1 (5.3%) | |
| **How often does use of Ophthalmology acronyms and/or Ophthalmology-specific terminology inhibit your understanding of an Ophthalmology note (i.e. APD, LPI, SLT, YAG)?** | | | 0.420 | | | 0.395 | | | 0.16 | | | 0.176 |
| Missing (N) | | | | | | | 0 | 1 | | | | |
| Never | 0 (0.0%) | 3 (8.1%) | | 2 (2.3%) | 2 (2.2%) | | 1 (2.1%) | 2 (5.4%) | | 1 (4.8%) | 2 (10.5%) | |
| Rarely | 1 (4.5%) | 1 (2.7%) | | 1 (1.1%) | 5 (5.5%) | | 0 (0.0%) | 4 (10.8%) | | 0 (0.0%) | 1 (5.3%) | |
| Sometimes | 2 (9.1%) | 7 (18.9%) | | 19 (21.8%) | 26 (28.6%) | | 13 (27.7%) | 9 (24.3%) | | 6 (28.6%) | 7 (36.8%) | |
| Often | 8 (36.4%) | 14 (37.8%) | | 43 (49.4%) | 39 (42.9%) | | 28 (59.6%) | 20 (54.1%) | | 9 (42.9%) | 9 (47.4%) | |
| Always | 11 (50.0%) | 12 (32.4%) | | 22 (25.3%) | 19 (20.9%) | | 5 (10.6%) | 2 (5.4%) | | 5 (23.8%) | 0 (0.0%) | |
| **Did the note improve your understanding of the patient's eye health?** | | | 0.139 | | | 0.236 | | | 0.373 | | | 0.42 |
| Missing (N) | | | | | | | 0 | 1 | | | | |
| Yes | 21 (95.5%) | 28 (75.7%) | | 76 (87.4%) | 71 (78.0%) | | 40 (85.1%) | 27 (73.0%) | | 16 (76.2%) | 17 (89.5%) | |
| No | 1 (4.5%) | 7 (18.9%) | | 8 (9.2%) | 16 (17.6%) | | 4 (8.5%) | 5 (13.5%) | | 4 (19.0%) | 1 (5.3%) | |
| I don't know | 0 (0.0%) | 2 (5.4%) | | 3 (3.4%) | 4 (4.4%) | | 3 (6.4%) | 5 (13.5%) | | 1 (4.8%) | 1 (5.3%) | |
| **How much did the note improve your understanding of this patient's Ophthalmology diagnosis(es)?*** | | | **0.047** | | | 0.389 | | | 0.105 | | | 0.587 |
| Not at all | 2 (9.1%) | 2 (5.4%) | | 5 (5.7%) | 6 (6.6%) | | 2 (4.3%) | 2 (5.3%) | | 2 (9.5%) | 1 (5.3%) | |
| Neutral | 0 (0.0%) | 7 (18.9%) | | 14 (16.1%) | 24 (26.4%) | | 8 (17.0%) | 15 (39.5%) | | 5 (23.8%) | 8 (42.1%) | |
| Moderately | 11 (50.0%) | 22 (59.5%) | | 37 (42.5%) | 33 (36.3%) | | 23 (48.9%) | 15 (39.5%) | | 10 (47.6%) | 6 (31.6%) | |

| | Resident/Fellow | | | Staff Physician | | | NP/PA | | | Allied Health | | |
|---|---|---|---|---|---|---|---|---|---|---|---|---|
| | Intervention (N=22) | Control (N=37) | p value | Intervention (N=87) | Control (N=91) | p value | Intervention (N=47) | Control (N=38) | p value | Intervention (N=21) | Control (N=19) | p value |
| A great deal | 9 (40.9%) | 6 (16.2%) | | 31 (35.6%) | 28 (30.8%) | | 14 (29.8%) | 6 (15.8%) | | 4 (19.0%) | 4 (21.1%) | |
| **After reading the entire note, were there any medical terms or concepts in the note that you found difficult to understand?*** | | | **0.023** | | | 0.321 | | | 0.677 | | | 0.997 |
| Missing (N) | 0 | 1 | | 2 | 1 | | | | | | | |
| Yes | 10 (45.5%) | 27 (75.0%) | | 30 (35.3%) | 39 (43.3%) | | 18 (38.3%) | 18 (47.4%) | | 9 (42.9%) | 8 (42.1%) | |
| No | 12 (54.5%) | 9 (25.0%) | | 55 (64.7%) | 50 (55.6%) | | 28 (59.6%) | 19 (50.0%) | | 11 (52.4%) | 10 (52.6%) | |
| I don't know | | | | 0 (0.0%) | 1 (1.1%) | | 1 (2.1%) | 1 (2.6%) | | 1 (4.8%) | 1 (5.3%) | |
| **How much did the note reduce your need to contact the consulting Ophthalmology team for clarification?** | | | 0.094 | | | 0.775 | | | 0.562 | | | 0.334 |
| Not at all | 5 (22.7%) | 6 (16.2%) | | 12 (13.8%) | 14 (15.4%) | | 4 (8.5%) | 4 (10.5%) | | 2 (9.5%) | 1 (5.3%) | |
| Neutral | 4 (18.2%) | 11 (29.7%) | | 35 (40.2%) | 41 (45.1%) | | 25 (53.2%) | 16 (42.1%) | | 7 (33.3%) | 11 (57.9%) | |
| Moderately | 2 (9.1%) | 11 (29.7%) | | 16 (18.4%) | 12 (13.2%) | | 10 (21.3%) | 7 (18.4%) | | 4 (19.0%) | 4 (21.1%) | |
| A great deal | 11 (50.0%) | 9 (24.3%) | | 24 (27.6%) | 24 (26.4%) | | 8 (17.0%) | 11 (28.9%) | | 8 (38.1%) | 3 (15.8%) | |
| **How satisfied or unsatisfied were you with the level of detail provided in the note?*** | | | **0.014** | | | **0.025** | | | **0.027** | | | 0.328 |
| Missing (N) | | | | 1 | 0 | | | | | | | |
| Not satisfied at all | 1 (4.5%) | 0 (0.0%) | | 2 (2.3%) | 0 (0.0%) | | 1 (2.1%) | 0 (0.0%) | | 0 (0.0%) | 0 (0.0%) | |
| Somewhat unsatisfied | 2 (9.1%) | 6 (16.2%) | | 8 (9.3%) | 8 (8.8%) | | 3 (6.4%) | 4 (10.5%) | | 2 (9.5%) | 1 (5.3%) | |
| Neutral | 0 (0.0%) | 3 (8.1%) | | 8 (9.3%) | 23 (25.3%) | | 5 (10.6%) | 14 (36.8%) | | 2 (9.5%) | 6 (31.6%) | |
| Somewhat satisfied | 3 (13.6%) | 16 (43.2%) | | 12 (14.0%) | 16 (17.6%) | | 9 (19.1%) | 7 (18.4%) | | 6 (28.6%) | 3 (15.8%) | |
| Satisfied | 16 (72.7%) | 12 (32.4%) | | 56 (65.1%) | 44 (48.4%) | | 29 (61.7%) | 13 (34.2%) | | 11 (52.4%) | 9 (47.4%) | |
| **How satisfied or unsatisfied were you with the conciseness of the note?** | | | 0.058 | | | 0.212 | | | 0.514 | | | 0.809 |
| Missing (N) | | | | 1 | 0 | | | | | | | |
| Not satisfied at all | 0 (0.0%) | 0 (0.0%) | | 0 (0.0%) | 0 (0.0%) | | 1 (2.1%) | 0 (0.0%) | | 0 (0.0%) | 0 (0.0%) | |
| Somewhat unsatisfied | 1 (4.5%) | 2 (5.4%) | | 4 (4.7%) | 1 (1.1%) | | 1 (2.1%) | 0 (0.0%) | | 2 (9.5%) | 1 (5.3%) | |
| Neutral | 0 (0.0%) | 4 (10.8%) | | 11 (12.8%) | 16 (17.6%) | | 5 (10.6%) | 7 (18.4%) | | 2 (9.5%) | 3 (15.8%) | |
| Somewhat satisfied | 1 (4.5%) | 9 (24.3%) | | 15 (17.4%) | 23 (25.3%) | | 7 (14.9%) | 8 (21.1%) | | 2 (9.5%) | 3 (15.8%) | |
| Satisfied | 20 (90.9%) | 22 (59.5%) | | 56 (65.1%) | 51 (56.0%) | | 33 (70.2%) | 23 (60.5%) | | 15 (71.4%) | 12 (63.2%) | |
| **How clear or unclear was the explanation of the patient's eye condition in the note?*** | | | **0.042** | | | **0.022** | | | **0.003** | | | 0.436 |
| Missing (N) | | | | 1 | 0 | | | | | | | |
| Not clear at all | 2 (9.1%) | 1 (2.7%) | | 5 (5.8%) | 2 (2.2%) | | 3 (6.4%) | 1 (2.6%) | | 1 (4.8%) | 1 (5.3%) | |
| Somewhat unclear | 1 (4.5%) | 7 (18.9%) | | 6 (7.0%) | 12 (13.2%) | | 1 (2.1%) | 6 (15.8%) | | 2 (9.5%) | 1 (5.3%) | |
| Neutral | 0 (0.0%) | 2 (5.4%) | | 5 (5.8%) | 9 (9.9%) | | 8 (17.0%) | 4 (10.5%) | | 1 (4.8%) | 5 (26.3%) | |

| | Resident/Fellow | | | Staff Physician | | | NP/PA | | | Allied Health | | |
|---|---|---|---|---|---|---|---|---|---|---|---|---|
| | Intervention (N=22) | Control (N=37) | p value | Intervention (N=87) | Control (N=91) | p value | Intervention (N=47) | Control (N=38) | p value | Intervention (N=21) | Control (N=19) | p value |
| Somewhat clear | 2 (9.1%) | 11 (29.7%) | | 15 (17.4%) | 29 (31.9%) | | 8 (17.0%) | 17 (44.7%) | | 6 (28.6%) | 4 (21.1%) | |
| Very clear | 17 (77.3%) | 16 (43.2%) | | 55 (64.0%) | 39 (42.9%) | | 27 (57.4%) | 10 (26.3%) | | 11 (52.4%) | 8 (42.1%) | |
| **If the patient were to ask a question about their ophthalmology diagnosis, eye health, or the general plan for ophthalmologic management, how confident would you be in your ability to answer such a question after reading the note?** | | | 0.206 | | | 0.415 | | | 0.737 | | | 0.820 |
| Missing (N) | | | | 1 | 0 | | | | | | | |
| Not comfortable | 0 (0.0%) | 3 (8.1%) | | 6 (7.0%) | 9 (9.9%) | | 2 (4.3%) | 3 (7.9%) | | 3 (14.3%) | 2 (10.5%) | |
| Somewhat uncomfortable | 4 (18.2%) | 8 (21.6%) | | 6 (7.0%) | 14 (15.4%) | | 4 (8.5%) | 6 (15.8%) | | 2 (9.5%) | 3 (15.8%) | |
| Neutral | 1 (4.5%) | 6 (16.2%) | | 7 (8.1%) | 6 (6.6%) | | 4 (8.5%) | 4 (10.5%) | | 4 (19.0%) | 6 (31.6%) | |
| Somewhat comfortable | 8 (36.4%) | 13 (35.1%) | | 30 (34.9%) | 28 (30.8%) | | 19 (40.4%) | 13 (34.2%) | | 5 (23.8%) | 3 (15.8%) | |
| Comfortable | 9 (40.9%) | 7 (18.9%) | | 37 (43.0%) | 34 (37.4%) | | 18 (38.3%) | 12 (31.6%) | | 7 (33.3%) | 5 (26.3%) | |
| ***Plain Language Summary vs Standard Note Questions*** | | | | | | | | | | | | |
| **Which version did you find easier to understand?** | | | | | | | | | | | | |
| Missing (N) | 2 | | | 9 | | | 2 | | | 1 | | |
| Standard Note, a great deal | 1 (5.0%) | | | 2 (2.6%) | | | 0 (0.0%) | | | 0 (0.0%) | | |
| Standard Note, somewhat | 1 (5.0%) | | | 3 (3.8%) | | | 1 (2.2%) | | | 1 (5.0%) | | |
| Neutral | 0 (0.0%) | | | 7 (9.0%) | | | 2 (4.4%) | | | 1 (5.0%) | | |
| Plain Language Summary, somewhat | 3 (15.0%) | | | 18 (23.1%) | | | 15 (33.3%) | | | 2 (10.0%) | | |
| Plain Language Summary, a great deal | 15 (75.0%) | | | 48 (61.5%) | | | 27 (60.0%) | | | 16 (80.0%) | | |
| **Which version provided clearer guidance regarding the patient's treatment plan?** | | | | | | | | | | | | |
| Missing (N) | 2 | | | 9 | | | 3 | | | 1 | | |
| Standard Note, a great deal | 1 (5.0%) | | | 3 (3.8%) | | | 0 (0.0%) | | | 0 (0.0%) | | |
| Standard Note, somewhat | 0 (0.0%) | | | 0 (0.0%) | | | 1 (2.3%) | | | 3 (15.0%) | | |
| Neutral | 2 (10.0%) | | | 6 (7.7%) | | | 5 (11.4%) | | | 4 (20.0%) | | |
| Plain Language Summary, somewhat | 5 (25.0%) | | | 23 (29.5%) | | | 12 (27.3%) | | | 0 (0.0%) | | |
| Plain Language Summary, a great deal | 12 (60.0%) | | | 46 (59.0%) | | | 26 (59.1%) | | | 13 (65.0%) | | |
| **Considering all aspects of your clinical practice, which type of note do you prefer?** | | | | | | | | | | | | |
| Missing (N) | 2 | | | 9 | | | 3 | | | 1 | | |
| Standard Note, a great deal | 1 (5.0%) | | | 2 (2.6%) | | | 0 (0.0%) | | | 0 (0.0%) | | |
| Standard Note, somewhat | 1 (5.0%) | | | 3 (3.8%) | | | 3 (6.8%) | | | 2 (10.0%) | | |
| Neutral | 0 (0.0%) | | | 8 (10.3%) | | | 2 (4.5%) | | | 3 (15.0%) | | |
| Plain Language Summary, somewhat | 4 (20.0%) | | | 18 (23.1%) | | | 11 (25.0%) | | | 0 (0.0%) | | |
| Plain Language Summary, a great deal | 14 (70.0%) | | | 47 (60.3%) | | | 28 (63.6%) | | | 15 (75.0%) | | |

**Supplementary Table 5.** BERTScore, SBERT, BLEU-4, and ROGUE-1 F-scores for PLSs compared to their corresponding SONs.

| ***Metric*** | | ***Mean*** | ***Std Dev*** | ***Min*** | ***Max*** |
|---|---|---|---|---|---|
| ***BERTScore*** | Precision | 0.85 | 0.04 | 0.71 | 0.95 |
| | Recall | 0.86 | 0.02 | 0.79 | 0.92 |
| | F1 Score | 0.85 | 0.03 | 0.75 | 0.94 |
| ***SBERT*** | Cosine Similarity | 0.84 | 0.06 | 0.57 | 0.98 |
| ***BLEU-4*** | | 0.08 | 0.07 | 0 | 0.48 |
| ***ROUGE-1*** | F1 Score | 0.47 | 0.11 | 0.12 | 0.81 |

**Supplementary Table 6.** Readability metrics of PLSs compared to readability metrics of SONs.

| | **Standard Ophthalmology Notes** | | | **Plain Language Summary** | | |
|---|---|---|---|---|---|---|
| ***Metric*** | *Mean +/- Std dev* | *Range (min, max)* | *Median (IQR)* | *Mean +/- Std dev* | *Range (min, max)* | *Median (IQR)* |
| ***Flesch Reading Ease*** | 41.65 +/- 19.21 | (-114.95,84.88) | 43.63 (19.83) | 50.9 +/- 9.23 | (17.34, 70.43) | 51.78 (11.21) |
| ***Flesch-Kincaid Grade Level*** | 13.64 +/- 6.75 | (4.4,77) | 11.9 (5.1) | 10.93 +/- 2.01 | (6.5, 22) | 10.7 (2.3) |
| ***SMOG Index*** | 12.17 +/- 5.05 | (0, 22.5) | 13.0 (3.6) | 13.08 +/- 1.49 | (9.3, 20.1) | 13.0 (1.9) |

IQR= Interquartile Range

### Supplementary Figures

Below is an instruction that describes a task. Write a response that appropriately completes the request. Your goal is to take this ophthalmology note and generate a plain language summary based on the Assessment and Plan where summary is easy to understand for someone who is not familiar with ophthalmology. Any ophthalmology term, diagnosis or finding MUST be followed by plain language translated. It is important to not use ophthalmology abbreviations. Refer to doctors as the Ophthalmology team. If there is a note with multiple dates just do the most recent date. It is important to ensure the summary does not change the original note's message. It is important you are correct. Keep the plan in a bullet point form for readability and ensure follow up is clear with the date if present. Ensure recommendations about follow up and treatment are included. Ensure all ophthalmology testing like 24-2 visual field testing, fluorescein angiography, and OCT is explained. Ensure timing of discharge testing is included.

**Supplementary Figure 1.** Prompt used to generate PLSs from SON text using LLM.

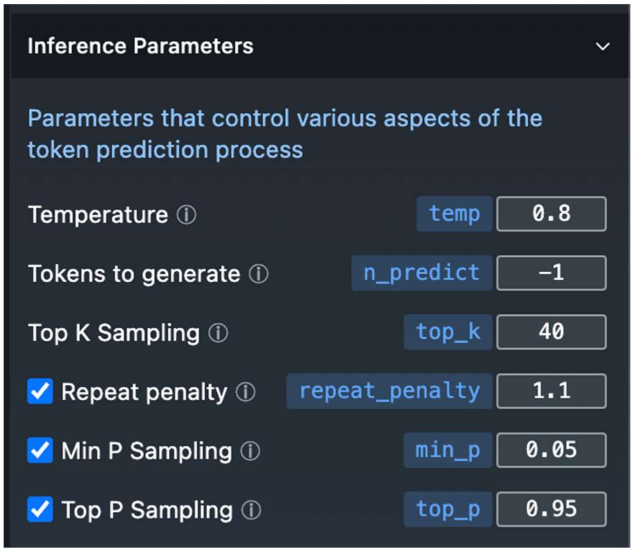

**Supplementary Figure 2.** LLM model parameters utilized for the duration of the study period.

----------------------------------------------------------------------------------------------------------------------------

----------------------------------------------------------------------------------------------------------------------------

As part of a Quality Improvement initiative to improve communication with the inpatient care team, we would like your feedback on the ophthalmology consult note. Please complete this quick 1-minute survey after reading the summary below. (Questions: Contact)
Survey: https://redcapcln-prod.mayo.edu/redcap/surveys/?s=JEFTMMYCK8TRWA8J
MRN: 5-122-792

**Supplementary Figure 3.** Text added to the end of each control arm SON including hyperlink to non-ophthalmologist survey.

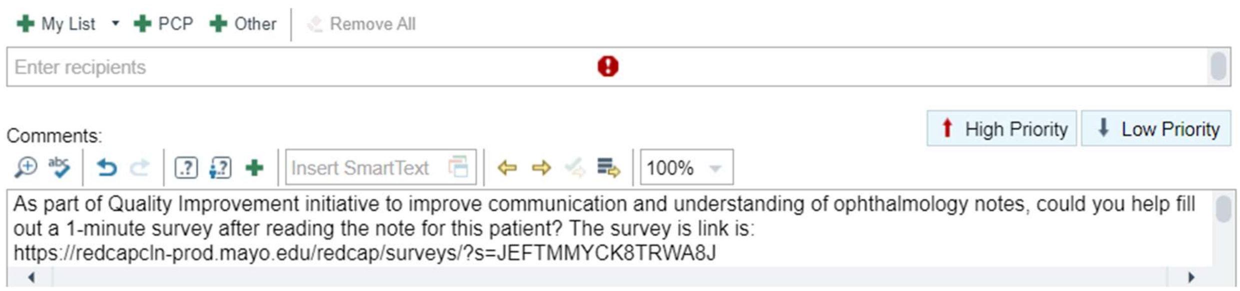

**Supplementary Figure 4.** Text sent as direct EMR message to non-ophthalmology care team members in both the control and intervention arm requesting participation in the survey.

**Plain Language Ophthalmology Summary (PLOS)**

Below is a Plain Language Ophthalmology Summary (PLOS) of the Ophthalmology consult, developed as part of our Quality Improvement initiative to improve communication with the care team. Your feedback is valuable; please complete this quick 1-minute survey after reading the summary below. (Questions: Contact)
Survey: https://redcapcln-prod.mayo.edu/redcap/surveys/?s=JEFTMMYCK8TRWA8J
MRN: 2-373-578

Summary:

**Supplementary Figure 5.** Text added to the end of each intervention arm SON prior to the PLS and including hyperlink to non-ophthalmologist survey.

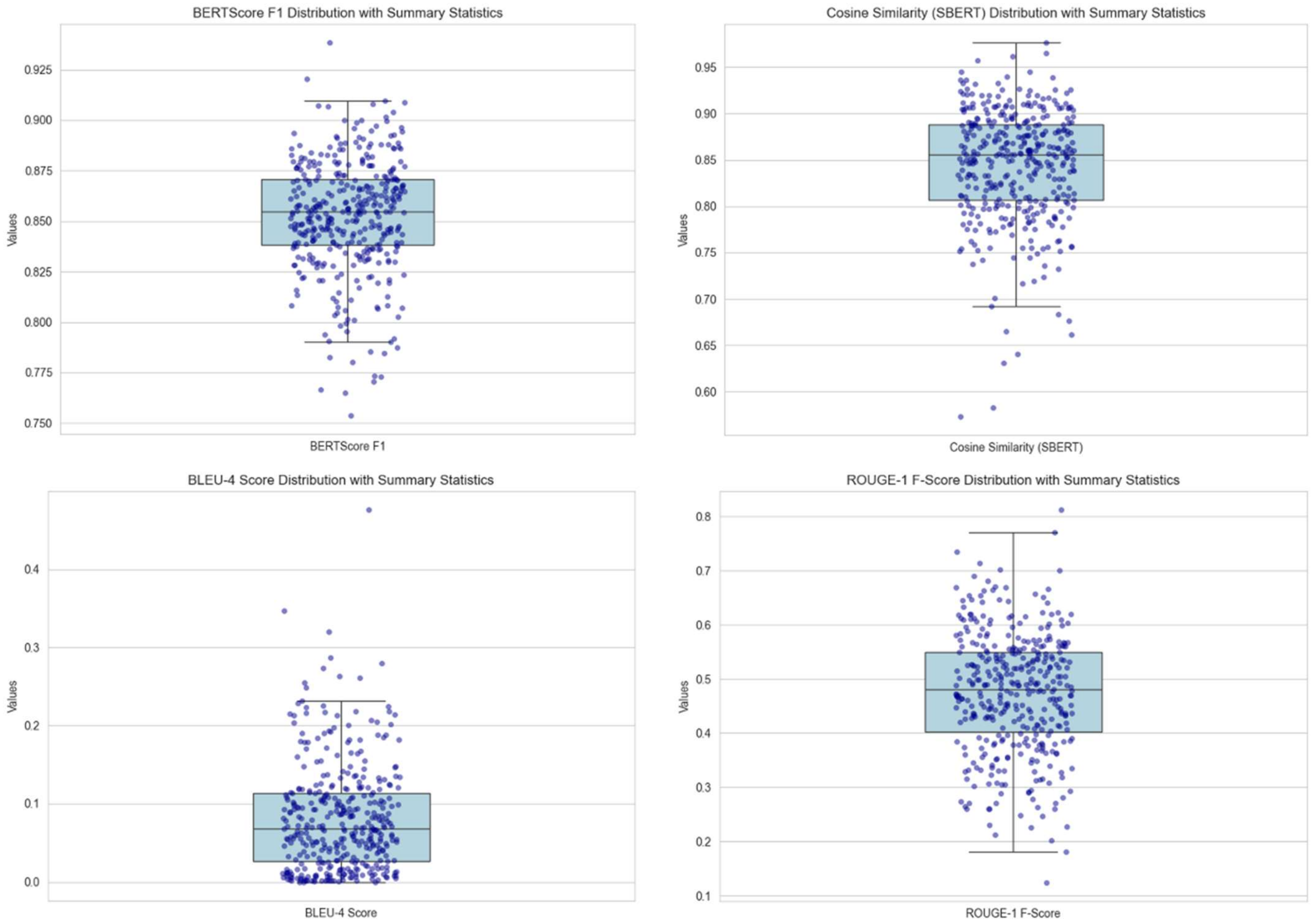

**Supplementary Figure 6.** Distribution of BERTScore, SBERT, BLEU-4, and ROGUE-1 F-scores for all PLSs compared to their corresponding SONs.
